# First-in-human trial evaluating safety and pharmacokinetics of AT-752, a novel nucleotide prodrug with pan-serotype activity against dengue virus

**DOI:** 10.1101/2024.01.03.24300771

**Authors:** Xiao-Jian Zhou, Jason Lickliter, Maureen Montrond, Laura Ishak, Keith Pietropaolo, Dayle James, Bruce Belanger, Arantxa Horga, Janet Hammond

**Affiliations:** Atea Pharmaceuticals, Inc, Boston, Massachusetts, USA; Nucleus Network, Melbourne, Australia

**Author notes:** Address correspondence to Xiao-Jian Zhou.

## Abstract

AT-752 is a novel guanosine nucleotide prodrug inhibitor of the dengue virus (DENV) polymerase with sub-micromolar, pan-serotype antiviral activity. This Phase 1, double-blind, placebo-controlled, first-in-human study evaluated the safety, tolerability, and pharmacokinetics of ascending single and multiple oral doses of AT-752 in healthy subjects. AT-752 was well tolerated when administered as a single dose up to 1500 mg, or when administered as multiple doses up to 750 mg three times daily (TID). No serious adverse events occurred, and the majority of treatment-emergent adverse events were mild in severity and resolved by the end of the study. In those receiving single ascending doses of AT-752, no pharmacokinetic ethnic sensitivity was observed in Asian subjects and no food effect was observed. Plasma exposure of the guanosine nucleoside metabolite AT-273, the surrogate of the active triphosphate metabolite of the drug, increased with increasing dose levels of AT-752 and exhibited a long half-life of approximately 15–25 h. Administration of AT-752 750 mg TID led to a rapid increase in plasma levels of AT-273 exceeding the target *in vitro* 90% effective concentration (EC_90_) of 0.64 μM in inhibiting DENV replication, and maintained this level over the treatment period. The favorable safety and pharmacokinetic results support evaluation of AT-752 as an antiviral for the treatment of dengue in future clinical studies.

## INTRODUCTION

Dengue is a mosquito-borne illness caused by four serotypes of dengue virus (DENV1–4) (1). DENV belongs to the Flaviviridae family and is a single-stranded positive-sense RNA virus (1). Dengue virus-associated diseases are major causes of illness and death in the tropical and subtropical world, with an estimated 100–400 million people infected yearly (https://www.who.int/news-room/fact-sheets/detail/dengue-and-severe-dengue). The incidence of dengue has increased 30-fold globally over the past 50 years, with several recent outbreaks contributing to a sharp increase in the Americas of over 3 million new infections recorded in 2023 (2, 3) (https://www.paho.org/en/news/3-8-2023-dengue-cases-increase-globally-vector-control-community-engagement-key-prevent-spread). Despite its high prevalence, there are no direct-acting antivirals for dengue, and treatment options are primarily centered around supportive care (4–6). There are currently two licensed dengue vaccines available in certain countries: CYD-TDV (chimeric yellow fever-dengue-tetravalent dengue vaccine; Dengvaxia) is licensed in 20 countries and indicated for use in individuals aged 9–45 years depending on country, whereas TAK-003 (Qdenga) is available for children and adults in Europe, Indonesia, Thailand, and Brazil (7–9) (https://www.takeda.com/newsroom/newsreleases/2023/Takeda-Dengue-Vaccine-ecommended-by-World-Health-Organization-Advisory-Group-for-Introduction-in-High-Dengue-Burden-and-Transmission-Areas-in-Children-Ages-Six-to-16-Years/). Critically, however, both vaccines offer imbalanced protection across serotypes, and data indicate low efficacy in younger children (7, 9, 10). Consequently, there are several vaccines and antivirals in development, including repurposed drugs (5–7). To date, clinical trials of repurposed drugs with antiviral activity such as balapiravir, chloroquine, lovastatin, and celgosivir have not demonstrated significant efficacy in reducing viremia and/or improvingclinical ou tcomes (11–14). Thus, there remains an unmet need for an effective direct-acting antiviral for the treatment of dengue, especially considering that a higher viral burden has been linked to severe dengue disease (6, 15).

AT-752 is an orally available novel guanosine nucleotide prodrug inhibitor of the DENV polymerase with sub-micromolar, pan-serotype antiviral activity (16). AT-752 is the hemisulfate salt of AT-281, a dual guanosine nucleotide prodrug which undergoes multistep metabolic activation to the active 5ill triphosphate metabolite AT-9010, which selectively inhibits the viral RNA-dependent RNA polymerase (16, 17) (**Figure 1**). Dephosphorylation of AT-9010 results in the formation of the guanosine nucleoside metabolite AT-273, which is regarded as a surrogate plasma marker for intracellular concentrations of AT-9010 (18).

**Figure 1.**
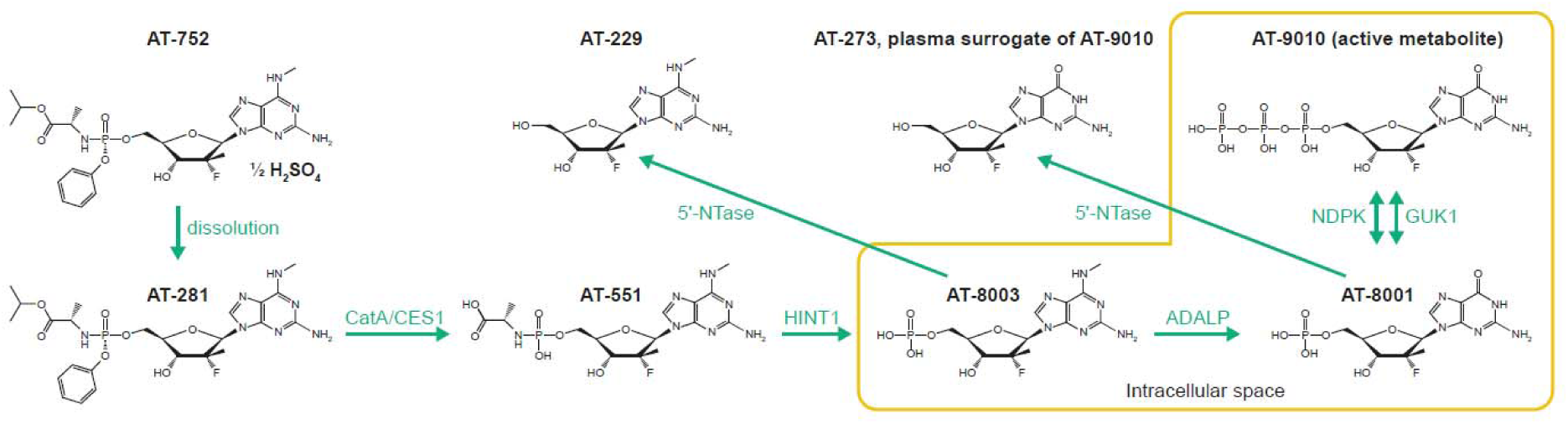
Metabolic pathway of AT-752. 5lil-NTase, 5lil-nucleotidase; ADALP, adenosine deaminase-like protein; CatA, cathepsin A; CES1, carboxylesterase 1; GUK1, guanylate kinase; HINT1, histidine triad nucleotide binding protein 1; NDPK, nucleoside-diphosphate kinase.

AT-752 is a potent inhibitor of DENV2 and DENV3, as well as other flaviviruses, *in vitro*, and demonstrated *in vivo* activity against DENV2 in a mouse model of disease (16). Based on initial pre-clinical toxicology studies of AT-752 in rats and monkeys, the no observed adverse effect level after 14 days of dosing was determined as 1000 mg/kg/day of AT-752 (expressed as AT-281 free base), providing at least 40-fold of margin of safety for the starting dose of 250 mg in humans (Data on file, Atea Pharmaceuticals, Inc).

This first-in-human Phase 1 study was designed to evaluate the safety, tolerability, and pharmacokinetics (PK) of oral single ascending doses (SAD) and multiple ascending doses (MAD) of AT-752 (free base AT-281 and metabolites AT-551, AT-229, and AT-273) in healthy male and female subjects. Additionally, the effect of food on the PK of single oral doses of AT-752, and PK ethnic sensitivity of AT-752 were assessed.

## RESULTS

### Study enrollment and completion

A total of 65 subjects were enrolled into one of six SAD or three MAD cohorts (see Materials and Methods). Among the SAD cohorts, 41 subjects were enrolled, and 40 subjects (97.6%) completed the trial. One subject (2.4%) randomized to receive AT-752 discontinued due to withdrawal of consent after receiving AT-752 500 mg (Cohort B), and was replaced. All 41 subjects were included in the safety population, and 31 subjects (75.6%) who received at least one dose of AT-752 were included in the PK population. Among the MAD cohorts, 24 subjects were enrolled and all completed the trial. All 24 subjects were included in the safety population, and 18 subjects (75.0%) who received at least one dose of AT-752 were included in the PK population.

### Patient demographics and baseline characteristics

Demographics and baseline characteristics in the SAD cohorts were generally similar across cohorts and treatments **(Table 1)**. The proportion of male (51.2%) and female (48.8%) subjects in the SAD cohorts were similar, whereas the majority were male (62.5%) in the

**Table 1.** Baseline characteristics of study subjects.

| Variable | SAD cohorts |  |  |  |  |  |  | MAD cohorts |  |  |  |  |
| --- | --- | --- | --- | --- | --- | --- | --- | --- | --- | --- | --- | --- |
|  | Cohort A<br>AT-752<br>250 mg<br>n=6 | Cohort B<br>AT-752<br>500 mg<br>n=7 | Cohort C<br>AT-752<br>1000 mg<br>n=6 | Cohort D<br>AT-752<br>1000 mg<br>ethnic<br>n=6 | Cohort E<br>AT-752<br>1500 mg<br>n=6 | Placebo<br>n=10 | Total<br>N=41 | Cohort F<br>AT-752<br>1000 mg QD<br>× 7 days<br>n=6 | Cohort G<br>AT-752<br>750 mg BID<br>× 4.5 days<br>n=6 | Cohort H<br>AT-752<br>750 mg TID<br>× 4.3 days<br>n=6 | Placebo<br>n=6 | Total<br>N=24 |
| Mean age, years<br>(range) | 28.2<br>(19–44) | 31.1<br>(16–64) | 28.5<br>(25–35) | 28.3<br>(21–43) | 30.8<br>(21–59) | 25.9<br>(20–39) | 28.6<br>(19–64) | 26.5<br>(20–33) | 38.5<br>(20–58) | 39.2<br>(21–62) | 37.2<br>(18–60) | 35.3<br>(18–62) |
| Sex, n (%) |  |  |  |  |  |  |  |  |  |  |  |  |
| Male | 3 (50.0) | 4 (57.1) | 2 (33.3) | 3 (50.0) | 3 (50.0) | 6 (60.0) | 21 (51.2) | 6 (100.0) | 3 (50.0) | 4 (66.7) | 2 (33.3) | 15 (62.5) |
| Female | 3 (50.0) | 3 (42.9) | 4 (66.7) | 3 (50.0) | 3 (50.0) | 4 (40.0) | 20 (48.8) | 0 | 3 (50.0) | 2 (33.3) | 4 (66.7) | 9 (37.5) |
| Race, n (%) |  |  |  |  |  |  |  |  |  |  |  |  |
| Asian | 3 (50.0) | 3 (42.9) | 2 (33.3) | 5 (83.3) | 3 (50.0) | 4 (40.0) | 20 (48.8) | 0 | 0 | 2 (33.3) | 0 | 2 (8.3) |
| White | 3 (50.0) | 4 (57.1) | 4 (66.7) | 0 | 3 (50.0) | 5 (50.0) | 19 (46.3) | 5 (83.3) | 6 (100.0) | 4 (66.7) | 6 (100.0) | 21 (87.5) |
| Other | 0 | 0 | 0 | 1 (16.7) | 0 | 1 (10.0) | 2 (4.9) | 1 (16.7) | 0 | 0 | 0 | 1 (4.2) |
| Mean height, cm<br>(range) | 169.8<br>(159–186) | 170.4<br>(162–178) | 167.7<br>(159–181) | 160.3<br>(143–172) | 175.2<br>(163–186) | 169.1<br>(162–187) | 168.8<br>(143–187) | 183.8<br>(177–187) | 174.8<br>(160–193) | 173.5<br>(161–187) | 166.0<br>(156–182) | 174.5<br>(156–193) |
| Mean weight, kg<br>(range) | 69.9<br>(51.9–79.9) | 64.2<br>(51.9–70.9) | 67.0<br>(51.9–77.8) | 58.6<br>(54.1–65.5) | 76.6<br>(59.4–94.0) | 67.5<br>(54.6–85.3) | 67.3<br>(51.9–94.0) | 77.5<br>(67.8–92.0) | 80.2<br>(66.3–95.9) | 73.2<br>(52.8–100.5) | 68.0<br>(60.6–89.0) | 74.7<br>(52.8–100.5) |

BID, twice daily; MAD, multiple ascending dose; QD, once daily; SAD, single ascending dose; TID, three times daily.

MAD cohorts. Overall, subjects were predominately white except for the SAD ethnic cohort where all participants were Asian.

### Safety and tolerability of AT-752

Overall, AT-752 was well tolerated in both the SAD and MAD cohorts, with no serious adverse events (SAEs) or discontinuations due to adverse events (AEs). Non-serious treatment-emergent adverse events (TEAEs) were mild or moderate in severity and resolved by the end of the study. The most frequently reported TEAEs are summarized in **Table 2**. Sporadic cases of gastrointestinal-related events, including mild-to-moderate vomiting, occurred mostly at higher doses. No treatment-related or dose-related trends in clinical laboratory values, vital sign measurements, or 12-lead electrocardiogram (ECG) parameters were observed.

**Table 2.** Summary of most frequent TEAEs reported by >2 subjects.

| TEAE | SAD cohorts |  |  |  |  |  |  |  | MAD cohorts |  |  |  |  | Overall<br>N=65 |
| --- | --- | --- | --- | --- | --- | --- | --- | --- | --- | --- | --- | --- | --- | --- |
|  | Cohort A<br>AT-752<br>250 mg<br>n=6 | Cohort B<br>AT-752<br>500 mg<br>n=7 | Cohort B<br>AT-752<br>500 mg<br>fed*<br>n=7 | Cohort C<br>AT-752<br>1000 mg<br>n=6 | Cohort D<br>AT-752<br>1000 mg<br>ethnic<br>n=6 | Cohort E<br>AT-752<br>1500 mg<br>n=6 | Pooled<br>placebo<br>n=10 | Total<br>N=41 | Cohort F<br>AT-752<br>1000 mg<br>QD × 7 days<br>n=6 | Cohort G<br>AT-752<br>750 mg BID<br>× 4.5 days<br>n=6 | Cohort H<br>AT-752<br>750 mg TID<br>× 4.3 days<br>n=6 | Pooled<br>placebo<br>n=6 | Total<br>N=24 |  |
| Headache,<br>n (%) | 0 | 2 (28.6) | 4 (57.1) | 1 (16.7) | 0 | 2 (33.3) | 1 (10.0) | 8 (19.5) | 0 | 1 (16.7) | 0 | 1 (16.7) | 2 (8.3) | 10 (15.4) |
| Nausea,<br>n (%) | 0 | 0 | 1 (14.3) | 1 (16.7) | 0 | 1 (16.7) | 0 | 3 (7.3) | 0 | 1 (16.7) | 0 | 1 (16.7) | 2 (8.3) | 5 (7.7) |
| Vomiting,<br>n (%) | 0 | 0 | 0 | 1 (16.7) | 0 | 2 (33.3) | 0 | 3 (7.3) | 0 | 1 (16.7) | 0 | 0 | 1 (4.2) | 4 (6.2) |

\*Food effect cohort.
BID, twice daily; QD, once daily; TEAE, treatment-emergent adverse event; TID, three times daily.

### PK evaluation

#### (i) SAD cohorts

Following single oral administration of AT-752 under fasting conditions, plasma concentrations of the parent prodrug AT-281 increased rapidly and were transient. AT-281 was rapidly eliminated with a short plasma half-life (t_1/2_) of approximately 0.5 h regardless of dose, resulting in mostly unquantifiable levels 4–6 h post dose. The intermediate L-alanyl metabolite AT-551 also exhibited a transient exposure with a mean t_1/2_ of approximately 2– 3 h across dose groups. The N^6^-methyl nucleoside metabolite, AT-229, subsequently peaked and exhibited a slower elimination phase (**Figure 2**). Plasma concentrations of AT-273 (surrogate for the intracellular active triphosphate AT-9010) appeared more gradually than AT-281 and other metabolites, and exhibited a long t_1/2_ of approximately 15–25 h across doses, reflecting sustained intracellular exposure of the active metabolite AT-9010.

**Figure 2.**
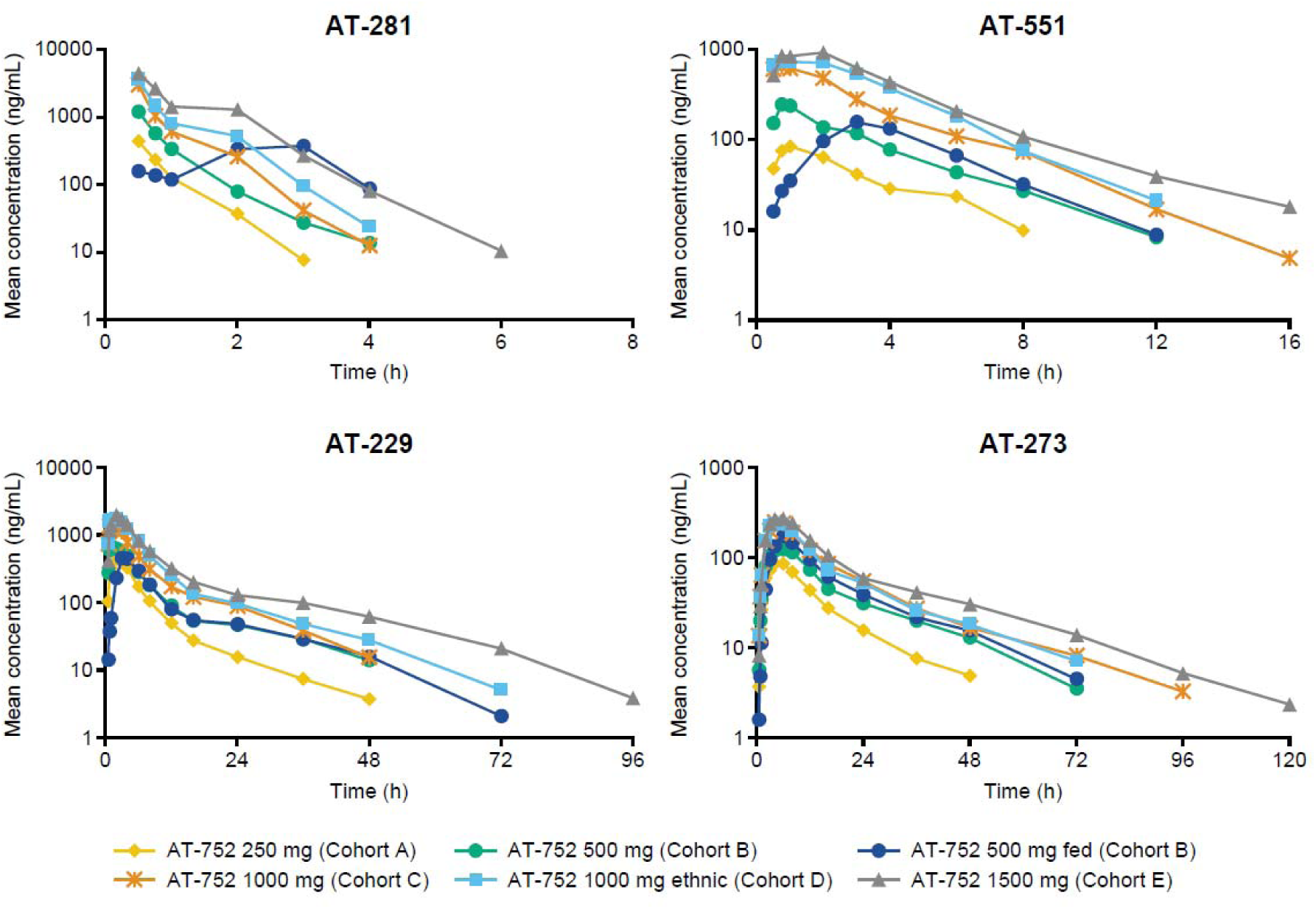
Mean plasma concentration–time profiles of AT-281 and its metabolites AT-551, AT-229, and AT-273 following oral administration of single ascending doses of AT-752.

Plasma PK parameters for the SAD cohorts are summarized in **Table 3**. Following single oral administration of AT-752 at 250–1500 mg, peak concentration (C_max_) and extent of exposure (AUC) were slightly higher numerically (with, however, largely overlapping ranges) for the ethnic cohort compared with the non-ethnic cohort at the same dose level (AT-752 1000 mg) for AT-281, AT-551, and AT-229. For AT-273, the guanosine circulating metabolite representing the intracellular active triphosphate metabolite AT-9010, C_max_, and AUC were similar for the ethnic cohort and the non-ethnic cohort at the same dose level. Time to peak concentration (T_max_) was also similar for the ethnic cohort and the non-ethnic cohort for all metabolites. The comparable PK profiles observed between the Asian and mostly Caucasian subject cohorts suggests an absence of PK ethnic sensitivity. Plasma exposure of AT-281 and its metabolites increased over the studied dose range of 250–1500 mg: AT-229 increased in a dose-proportional manner, and AT-281 and AT-551 increased in a greater than dose-proportional manner, whereas AT-273 was slightly less than dose-proportional (see Figure S1 in the Supplementary Materials). A high-fat/high-calorie meal delayed and decreased peak levels of AT-281, AT-551, and AT-229 but had limited-to-no impact on their total exposure, and slightly increased the plasma exposure of AT-273 in comparison to fasted conditions. This demonstrates that AT-752 can therefore be taken with or without food. The extent of urine elimination was low for AT-281 (up to 2.4%) and AT-551 (up to 0.7%), and moderate for AT-229 (10.7–17.9%) and AT-273 (6.7–8.8%). Total urine recovery ranged from approximately 20–30% of administered doses across cohorts, and renal clearance (CL_R_) of AT-229 and AT-273 exceeded estimated glomerular filtration rate (eGFR).

**Table 3.** Summary of plasma PK parameters of AT-281 and metabolites AT-551, AT-229, and AT-273 following oral administration of single ascending doses of AT-752.

| Cohort | C<sub>max</sub><br>(ng/mL) | T<sub>max</sub><br>(h) | AUC<sub>inf</sub><br>(ng/mL*h) | t<sub>1/2</sub><br>(h) | CL<sub>R</sub><br>(L/h) |
| --- | --- | --- | --- | --- | --- |
| **AT-281** |  |  |  |  |  |
| 250 mg (Cohort A) | 482±131 | 0.5 (0.5–0.8) | 347±64.0 | 0.5±0.1 | 7.5±1.5 |
| 500 mg (Cohort B) | 1230±1110 | 0.5 (0.5–6.0) | 1110±687 | 0.5±0.1 | 10.9±5.4 |
| 500 mg fed (Cohort B)<br>Food effect* | 488±303<br>52.2 (22.1–123.6) | 2.0 (0.5–3.0) | 980±716<br>80.2 (38.7–166.2) | 0.7±0.3 | 6.6±1.6 |
| 1000 mg (Cohort C) | 3040±960 | 0.5 (0.5–0.6) | 1910±508 | 0.5±0.1 | 11.4±1.7 |
| 1000 mg ethnic (Cohort D) | 3670±1260 | 0.5 (0.5–0.5) | 2730±944 | 0.5±0.1 | 9.7±4.2 |
| 1500 mg (Cohort E) | 5050±2080 | 0.5 (0.5–0.8) | 4630±1500 | 0.6±0.2 | 8.3±2.2 |
| **AT-551** |  |  |  |  |  |
| 250 mg (Cohort A) | 89.2±35.6 | 1.0 (0.8–2.0) | 302±70.5 | 2.1±0.3 | 2.0±0.3 |
| 500 mg (Cohort B) | 231±155 | 1.0 (0.8–8.0) | 898±437 | 2.4±0.5 | 2.1±0.6 |
| 500 mg fed (Cohort B)<br>Food effect* | 154±83.6<br>70.5 (42.8–116.0) | 3.1 (3.0–4.0) | 737±289<br>78.3 (61.8–99.2) | 2.3±0.4 | 2.1±0.4 |
| 1000 mg (Cohort C) | 695±468 | 0.8 (0.5–2.0) | 2330±1060 | 2.2±0.4 | 2.1±0.6 |
| 1000 mg ethnic (Cohort D) | 895±418 | 0.9 (0.5–2.0) | 3320±1630 | 2.0±0.3 | 2.1±0.6 |
| 1500 mg (Cohort E) | 1080±217 | 1.5 (0.8–2.0) | 4100±494 | 3.2±1.2 | 1.9±0.4 |
| **AT-229** |  |  |  |  |  |
| 250 mg (Cohort A) | 446±121 | 2.1 (2.0–3.0) | 2890±680 | 16.9±8.6 | 13.7±1.6 |
| 500 mg (Cohort B) | 765±307 | 1.0 (0.8–4.0) | 5480±1630 | 15.1±6.2 | 13.6±5.4 |
| 500 mg fed (Cohort B)<br>Food effect* | 488±324<br>64.9 (43.8–96.2) | 4.0 (3.0–6.1) | 4310±1400<br>78.4 (68.4–89.9) | 11.2±4.9 | 12.5±2.8 |
| 1000 mg (Cohort C) | 1640±474 | 0.9 (0.8–3.0) | 10300±3070 | 12.4±10.7 | 14.2±3.8 |
| 1000 mg ethnic (Cohort D) | 2010±489 | 1.5 (0.8–2.0) | 14000±4210 | 14.7±10.1 | 13.4±2.5 |
| 1500 mg (Cohort E) | 2030±600 | 2.0 (1.0–2.0) | 16700±3780 | 12.5±3.8 | 15.0±2.4 |
| **AT-273** |  |  |  |  |  |
| 250 mg (Cohort A) | 90.8±18.9 | 6.0 (3.0–6.0) | 1440±280 | 19.1±10.2 | 15.8±2.8 |
| 500 mg (Cohort B) | 135±24.9 | 4.0 (3.0–8.0) | 2390±478 | 13.9±5.9 | 16.8±7.6 |
| 500 mg fed (Cohort B)<br>Food effect* | 175±28.8<br>132.0 (109.8–158.6) | 6.0 (6.0–8.1) | 2990±558<br>125.6 (111.2–141.8) | 17.3±11.0 | 13.4±3.3 |
| 1000 mg (Cohort C) | 241±71.1 | 4.1 (4.0–6.3) | 4350±1520 | 26.2±12.2 | 17.1±6.5 |
| 1000 mg ethnic (Cohort D) | 240±46.2 | 5.0 (3.1–6.0) | 3830±519 | 16.3±3.0 | 18.7±2.1 |
| 1500 mg (Cohort E) | 283±31.6 | 6.0 (3.2–6.1) | 5460±1190 | 21.7±6.2 | 19.5±4.1 |

\*Food effect is represented as geometric mean ratio (90% CI).
C<sub>max</sub>, AUC<sub>inf</sub>, and t<sub>1/2</sub> are represented as mean ± SD. T<sub>max</sub> is represented as median (minimum–maximum). CL<sub>R</sub> is represented as mean ± SD.
AUC<sub>inf</sub>, area under the curve extrapolated to infinity; CI, confidence interval; CL<sub>R</sub>, renal clearance; C<sub>max</sub>, maximum plasma concentration; SD, standard deviation; t<sub>1/2</sub>, half-life; T<sub>max</sub>, time to reach maximum concentration.

#### (ii) MAD cohorts

Following repeat dosing of AT-752 1000 mg once daily (QD), 750 mg twice daily (BID), and 750 mg three times daily (TID), plasma AT-281 (the unchanged protide) did not meaningfully accumulate due to its rapid elimination (**Figure 3**). Plasma exposure of AT-273 increased by approximately 25, 60, and 80% with QD, BID, and TID, respectively, due to its long plasma half-life, reflecting rapid accumulation and sustained intracellular exposure of the active metabolite AT-9010. Steady-state plasma PK parameters are summarized in **Table 4**. Based on the trough AT-273 concentrations, steady state was essentially achieved by Day 3 in all MAD cohorts (**Figure 4**).

**Figure 3.**
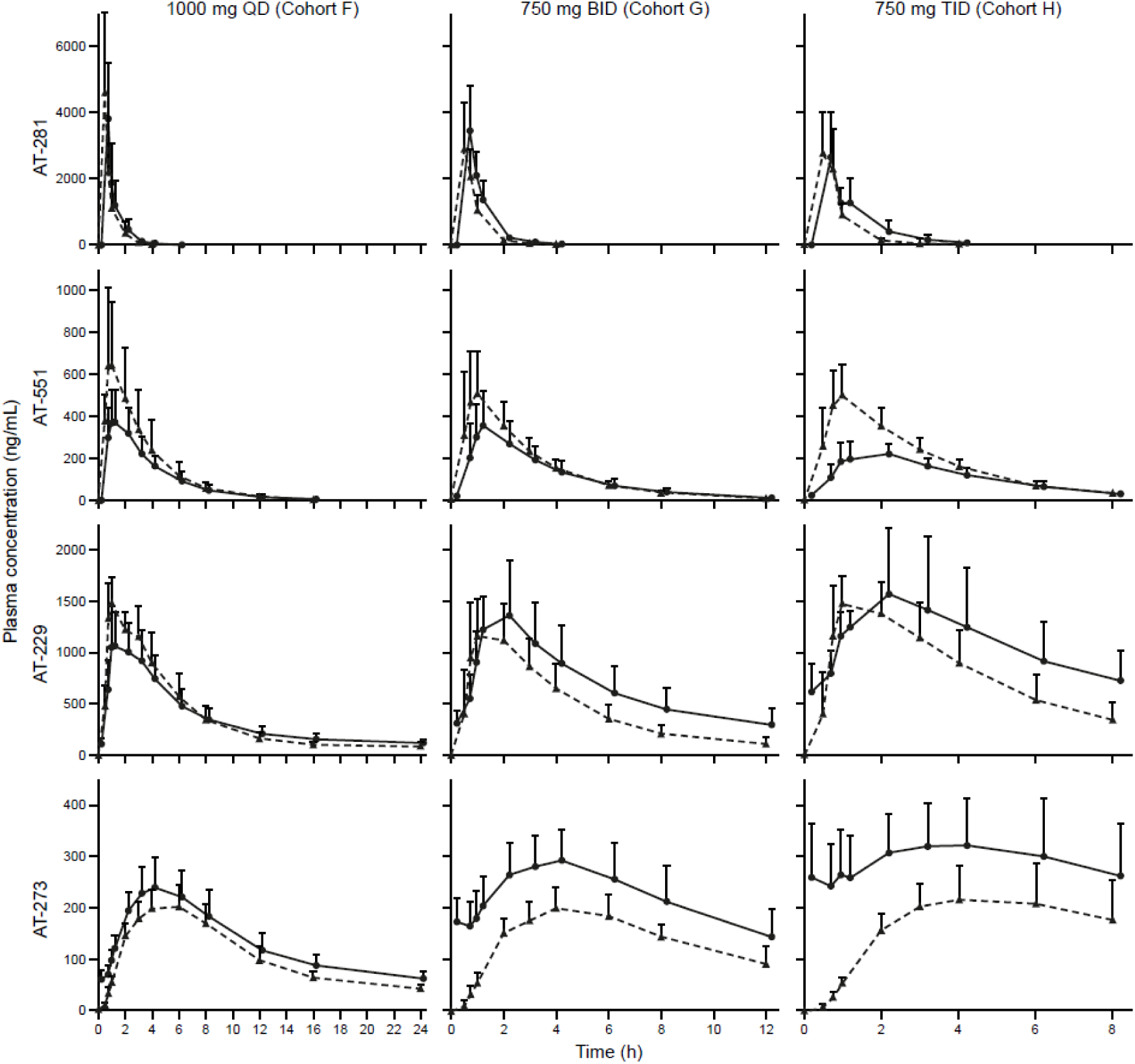
Mean (+ SD) plasma concentration–time profiles of AT-281 and metabolites AT-551, AT-229, and AT-273 following oral administration of multiple ascending doses of AT-752. Dashed line: first dose on Day 1; solid line: last dose at steady state. BID, twice daily; QD, once daily; SD, standard deviation; TID, three times daily.

**Figure 4.**
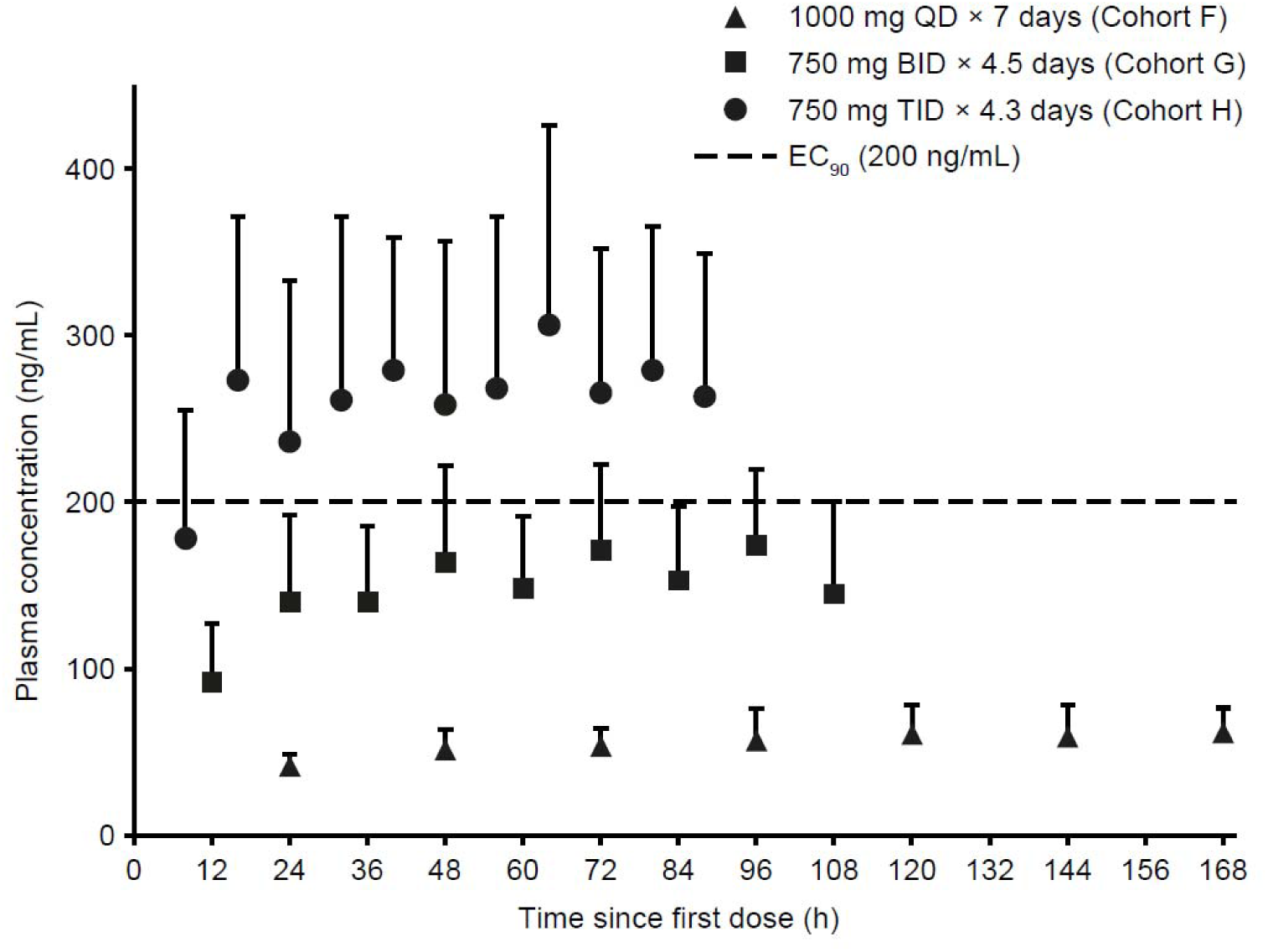
Mean (+ SD) plasma trough concentration–time profiles of AT-273 with following oral administration of multiple ascending doses of AT-752. BID, twice daily; EC_90_, 90% effective concentration; QD, once daily; SD, standard deviation; TID, three times daily.

**Table 4.** Summary of steady-state plasma PK parameters of AT-281 and metabolites AT-551, AT-229, and AT-273 following oral administration of multiple ascending doses of AT-752.

| Cohort | C<sub>max</sub><br>(ng/mL) | T<sub>max</sub><br>(h) | AUC<sub>tau</sub><br>(ng/mL*h) |
| --- | --- | --- | --- |
| **AT-281** |  |  |  |
| 1000 mg QD (Cohort F) | 3810±1670 | 0.5 (0.5–0.5) | 3090±1530 |
| 750 mg BID (Cohort G) | 3490±1280 | 0.5 (0.5–0.8) | 2690±640 |
| 750 mg TID (Cohort H) | 3010±754 | 0.5 (0.5–1.0) | 2540±572 |
| **AT-551** |  |  |  |
| 1000 mg QD (Cohort F) | 392±143 | 1.0 (0.8–2.0) | 1650±498 |
| 750 mg BID (Cohort G) | 365±159 | 1.0 (0.8–2.0) | 1330±503 |
| 750 mg TID (Cohort H) | 245±30.2 | 1.5 (0.8–2.0) | 944±173 |
| **AT-229** |  |  |  |
| 1000 mg QD (Cohort F) | 1200±284 | 0.9 (0.8–2.0) | 8290±2290 |
| 750 mg BID (Cohort G) | 1470±404 | 1.5 (0.8–2.2) | 7960±2970 |
| 750 mg TID (Cohort H) | 1600±640 | 1.5 (1.0–2.0) | 8880±3330 |
| **AT-273** |  |  |  |
| 1000 mg QD (Cohort F) | 241±56.3 | 4.0 (3.0–4.0) | 3120±694 |
| 750 mg BID (Cohort G) | 288±58.4 | 3.5 (3.0–4.0) | 2680±730 |
| 750 mg TID (Cohort H) | 330±97.7 | 3.5 (3.0–6.0) | 2370±743 |

C<sub>max</sub> and AUC<sub>tau</sub> represented as mean ± SD. T<sub>max</sub> is represented as median (minimum–maximum).
AUC<sub>tau</sub>, area under plasma concentration-time curve over the dosing interval; BID, twice daily; C<sub>max</sub>, peak concentration; PK, pharmacokinetic; QD, once daily; TID, three times daily; T<sub>max</sub>, time to peak concentration.

As depicted in Figure 4, among the MAD cohorts, only AT-752 750 mg TID led to a rapid increase in plasma AT-273 levels, with mean trough levels within the first day exceeding the 90% effective concentration (EC_90_) of the drug in inhibiting DENV replication in vitro (0.64 μM or 200 ng/mL of equivalent AT-273), and maintained these levels over the treatment period.

## DISCUSSION

AT-752, when metabolized to the active triphosphate AT-9010, has been demonstrated to target the NS5 RNA-dependent RNA polymerase (RdRp) of DENV1–4 (17). NS5, the largest and most conserved nonstructural protein encoded by flaviviruses, is a promising target for antivirals, particularly nucleoside/nucleotide analogs in the treatment of dengue (5). This first-in-human Phase 1 study of AT-752 evaluated the safety, tolerability, and PK of ascending single and multiple doses in healthy subjects. AT-752 demonstrated a favorable safety profile and was well tolerated in healthy subjects when administered as a single oral dose up to 1500 mg, or when administered as multiple oral doses up to 750 mg TID. No SAEs were observed, and no subjects experienced AEs that lead to study drug discontinuation. Most TEAEs were mild in severity, and all TEAEs resolved by the end of the study. There were no clinically significant physical examination findings, vital signs, or ECGs observed.

In the SAD cohorts, plasma exposure of AT-281 and its metabolites was approximately dose-proportional over the studied dose range of 250–1500 mg AT-752. Fed conditions led to a 48% reduction in C_max_ compared with fasted conditions, whereas AUC values were similar. The extent of urinary elimination of AT-281 and its metabolites was low-to-moderate. Renal clearance of the nucleoside metabolites AT-229 and AT-273 exceeded eGFR, suggesting involvement of active secretion in their renal elimination. Furthermore, the similar PK profiles observed between the ethnic and non-ethnic cohorts suggests an absence of PK ethnic sensitivity in Asian participants; therefore, dose adjustment of AT-752 in this population does not appear necessary. In the MAD cohorts, C_max_ of AT-281 were attained approximately 0.5 h post dose (median estimates) for all dose levels. Plasma exposure of AT-273 increased with increasing dose levels due to its long plasma half-life, reflecting rapid accumulation and sustained intracellular exposure of the active metabolite AT-9010. AT-752 750 mg TID led to a rapid increase in plasma AT-273 levels, exceeding the target EC_90_ of 0.64 μM, and maintained this level over the treatment period.

In summary, the results of this study demonstrate that AT-752 was well tolerated when administered as a single dose up to 1500 mg, or when administered as multiple doses up to 750 mg TID. In the SAD cohorts, AT-752 exhibited no PK ethnic sensitivity in South/Southeast/East Asian participants and no food effect was observed, demonstrating that ethnic dose adjustment is not necessary and that AT-752 can be taken with or without food. In the MAD cohorts, AT-752 TID exceeded the target antiviral level for inhibition of DENV replication. The favorable safety and PK results reported here demonstrate AT-752 as an attractive antiviral for the treatment of dengue, and support dose selection of AT-752 in future clinical studies.

## MATERIALS AND METHODS

The study protocol was approved by the Human Research Ethics Committee before the trial began, and written informed consent was obtained from each subject before entering the study. This study was conducted according to the principles of the International Council for Harmonisation harmonised tripartite guideline E6(R2): Good Clinical Practice, and the ethical principles from the Declaration of Helsinki.

### Study design

This was a Phase 1, first-in-human, randomized, double-blind, placebo-controlled study consisting of two sequential parts: SAD and MAD. The SAD cohorts included a food-effect cohort and both SAD and MAD cohorts evaluated the safety, tolerability, and PK of AT-752 (ClinicalTrials.gov registration no. NCT04722627). All subjects were screened up to 28 days prior to the first study drug administration on Day 1, which included clinical history, physical examination, 12-lead ECG, vital signs, and laboratory tests of blood and urine.

#### (i) SAD cohorts

Subjects in the SAD cohorts were assigned to one of five sequential dose cohorts (eight subjects per cohort) on Day 1 prior to the first dose. Subjects were randomized 3:1 within each cohort to receive AT-752 or a matching placebo (**Table 5**). Sentinel dosing was employed for two subjects in each SAD cohort in a ratio of 1:1 (one active, one placebo), and were dosed at least 48 h before the remainder of the cohort was dosed. All treatments were administered orally under fasting conditions, except for the food effect cohort. As part of the food effect evaluation, subjects in Cohort B (n=7) remained confined to the clinic for an additional study drug administration on Day 7 under fed conditions. Subjects fasted overnight for at least 8 h and received a high-fat and high-calorie breakfast approximately 30 minutes (± 5 minutes) before study drug administration. Subjects consumed the meal within 30 minutes or less and consumed at least 75% of the meal. The high-fat (approximately 50% of total calorific content of the meal) and high-calorie (approximately 800–1000 calories) breakfast, followed Food and Drug Administration (FDA) guidance recommendations and provided approximately 150, 250, and 500–600 calories from protein, carbohydrate, and fat, respectively.

**Table 5.** SAD cohorts.

| Cohort | Dose (mg) | Administration |
| --- | --- | --- |
| A | 250 | Day 1 (fasting) |
| B (+ food effect) | 500 | Day 1 (fasting); Day 7 (fed) |
| C | 1000 | Day 1 (fasting) |
| D (Ethnic) | 1000 | Day 1 (fasting) |
| E | 1500 | Day 1 (fasting) |

#### (ii) MAD cohorts

Subjects in the MAD cohorts were assigned to a repeat-dose cohort on Day 1 prior to the first dose. Subjects were randomized 3:1 to receive AT-752 or a matching placebo (**Table 6**). Sentinel dosing was employed for two subjects in each MAD cohort in a ratio of 1:1 (one active, one placebo), and were dosed at least 48 h before the remainder of the cohort was dosed. All treatments were administered orally under fasting conditions. Dose escalation to Cohort G occurred based on review of available safety data (through Day 12) and PK data (through Day 8) of Cohort F. Similarly, dose escalation to Cohort H occurred based on review of available safety data (through Day 10) and PK data (through Day 6) of Cohort G.

**Table 6.**
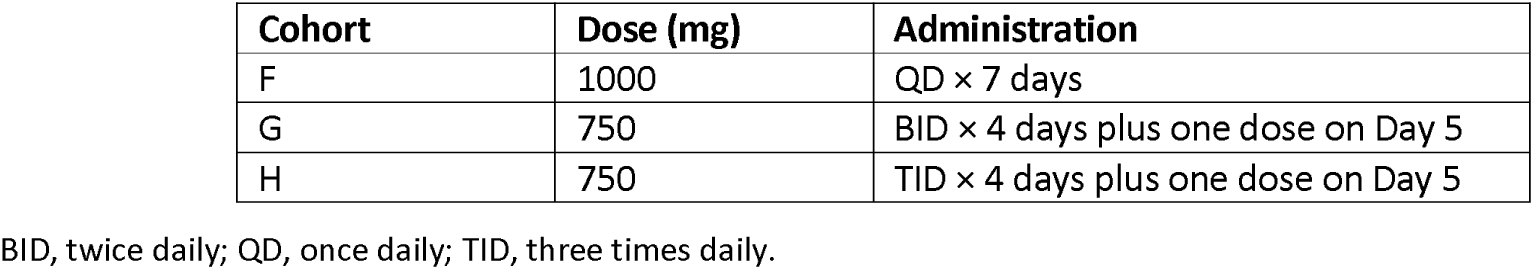
MAD cohorts.

### Study subjects

Healthy subjects between the ages of 18 and 65 years who met the inclusion and exclusion criteria were considered eligible for this study. All subjects were required to weigh at least 50 kg and have a body mass index of between 18 and 29 kg/m, inclusive. Subjects in Cohort D (ethnic cohort) must have been of first to third generation South/Southeast/East Asian descent. Exclusion criteria included clinically relevant abnormal medical history, physical findings, ECG, or laboratory values at the pre-trial screening assessment; positive test for hepatitis B surface antigen, hepatitis C virus antibody, human immunodeficiency virus types 1 or 2 antibodies, or SARS-CoV-2 at screening; or receipt of a vaccine against COVID-19 in the 14 days before the first dose. Subjects could not have used a prescription medicine, over-the-counter medicine, or herbal or dietary supplements during the 7 days before the first dose.

### Pharmacokinetic assessments

Blood samples for plasma PK analyses were collected pre dose and 0.5, 0.75, 1, 2, 3, 4, 6, 8, 12, 16, 24, 36, 48, 72, 96, and 120 h post dose for the SAD cohorts. For the food effect cohort, blood samples were also taken on Day 7 (fed state) at ≤5 minutes prior to dosing and 0.5, 0.75, 1, 2, 3, 4, 6, 8, 12, 16, 24, 36, 48, 72, 96, and 120 h post dose. For Cohort F, blood samples were collected prior to dosing and 0.5, 0.75, 1, 2, 3, 4, 6, 8, 12, and 16 h post dose on Day 1; ≤5 minutes prior to dosing and 4 h post dose on Days 2–6; ≤5 minutes prior to dosing and 0.5, 0.75, 1, 2, 3, 4, 6, 8, 12, 16, 24, 36, 48, 72, 96, and 120 h post dose on Day 7. For Cohort G, blood samples were collected prior to dosing and 0.5, 0.75, 1, 2, 3, 4, 6, and 8 h post dose on Day 1 (AM); ≤5 minutes prior to dosing on Day 1 (PM); ≤5 minutes prior to AM and PM dosing on Days 2–4; ≤5 minutes prior to AM dosing and 0.5, 0.75, 1, 2, 3, 4, 6, 8, 12, 16, 24, 36, 48, 72, 96, and 120 h post dose on Day 5. For Cohort H, blood samples were collected prior to dosing and 0.5, 0.75, 1, 2, 3, 4, and 6 h post dose on Day 1 (AM); ≤5 minutes prior to PM dosing on Day 1; ≤5 minutes prior to AM and PM dosing on Days 2 and 3; ≤5 minutes prior to AM dosing (Day 4); ≤5 minutes prior to PM dosing and 0.5, 0.75, 1, 2, 3, 4, and 6 h post dose on Day 4 (PM); ≤5 minutes prior to AM dosing and 0.5, 0.75, 1, 2, 3, 4, 6, 8, 12, 16, 24, 48, 72, 96, and 120 h post dose on Day 5. Urine samples for PK analysis were collected prior to dosing and 0–4, 4–8, 8–12, 12–24, 24–48, 48–72, 72–96, and 96–120 h post dose on Day 1 for the SAD cohorts, as well as on Day 7 (fed state) for the food effect cohort. For Cohort F, urine samples were collected prior to dosing on Day 1, and a pooled sample taken over 0–24 h post dose on Day 7. The collected plasma and urine samples underwent quantitation for AT-281 and its metabolites AT-551, AT-229, and AT-273 using validated liquid chromatography/tandem mass spectrometry (LC-MS/MS) methodologies. Plasma assay ranges were 5–5000 ng/mL for AT-281 and AT-551, and 2–2000 ng/mL for AT-229 and AT-273; urine assay ranges were 5–5000 ng/mL for AT-281 and AT-551, and 10–10000 ng/mL for AT-229 and AT-273.

### Statistical analysis

Plasma concentration–time data were analyzed by non-compartmental approach using Phoenix® WinNonlin® Version 8.3 (Certara USA Inc., Princeton, New Jersey) or SAS® Version 9.4 (SAS Institute Inc., Cary, North Carolina), as appropriate. For the SAD cohorts, dose proportionality was assessed on Day 1 for AT-281, AT-551, AT-229, and AT-273 in the plasma using the power regression model for the AUCs and C_max_ PK parameters (19).

To assess the impact of a high-fat meal on exposure to AT-281, AT-551, AT-229, and AT-273, the log transformed values of AUC_inf_ and C_max_ were analyzed in Cohort B for AT-281, AT-551, AT-229, and AT-273 using a linear mixed effect model with fed/fasted status as a fixed effect and subject as a random effect.

Ethnic sensitivity analysis for analyte AT-752 PK was performed by visual inspection of mean concentration vs time profiles and assessment of PK parameters in summary tables.

### Laboratory and safety assessment

Safety assessments included monitoring of AEs, clinical laboratory tests (hematology, serum chemistry, urinalysis, coagulation tests, and cardiac biomarkers), vital sign measurements, 12-lead ECGs, and physical examination. All AEs were coded using Medical Dictionary for Regulatory Activities (MedDRA) Version 24.1.

## Data Availability

All data produced in the present study are available upon reasonable request to the authors

## ACKNOWLEDGEMENTS

This study was funded by Atea Pharmaceuticals, Inc. Medical writing support was provided by Samantha Brick, Elements Communications Ltd, UK, and funded by Atea Pharmaceuticals, Inc.

Xiao-Jian Zhou, Maureen Montrond, Laura Ishak, Keith Pietropaolo, Dayle James, Bruce Belanger, Arantxa Horga, Janet Hammond are employees of and may own stock in Atea Pharmaceuticals, Inc, Boston, MA, USA; Jason Lickliter is an employee of Nucleus Network, Melbourne, Australia, which was contracted by Atea Pharmaceuticals, Inc to help perform this research.

