## Supplemental Figure 1 for "First-in-human trial evaluating safety and pharmacokinetics of AT-752, a novel nucleotide prodrug with pan-serotype activity against dengue virus"

**Supplementary Materials**

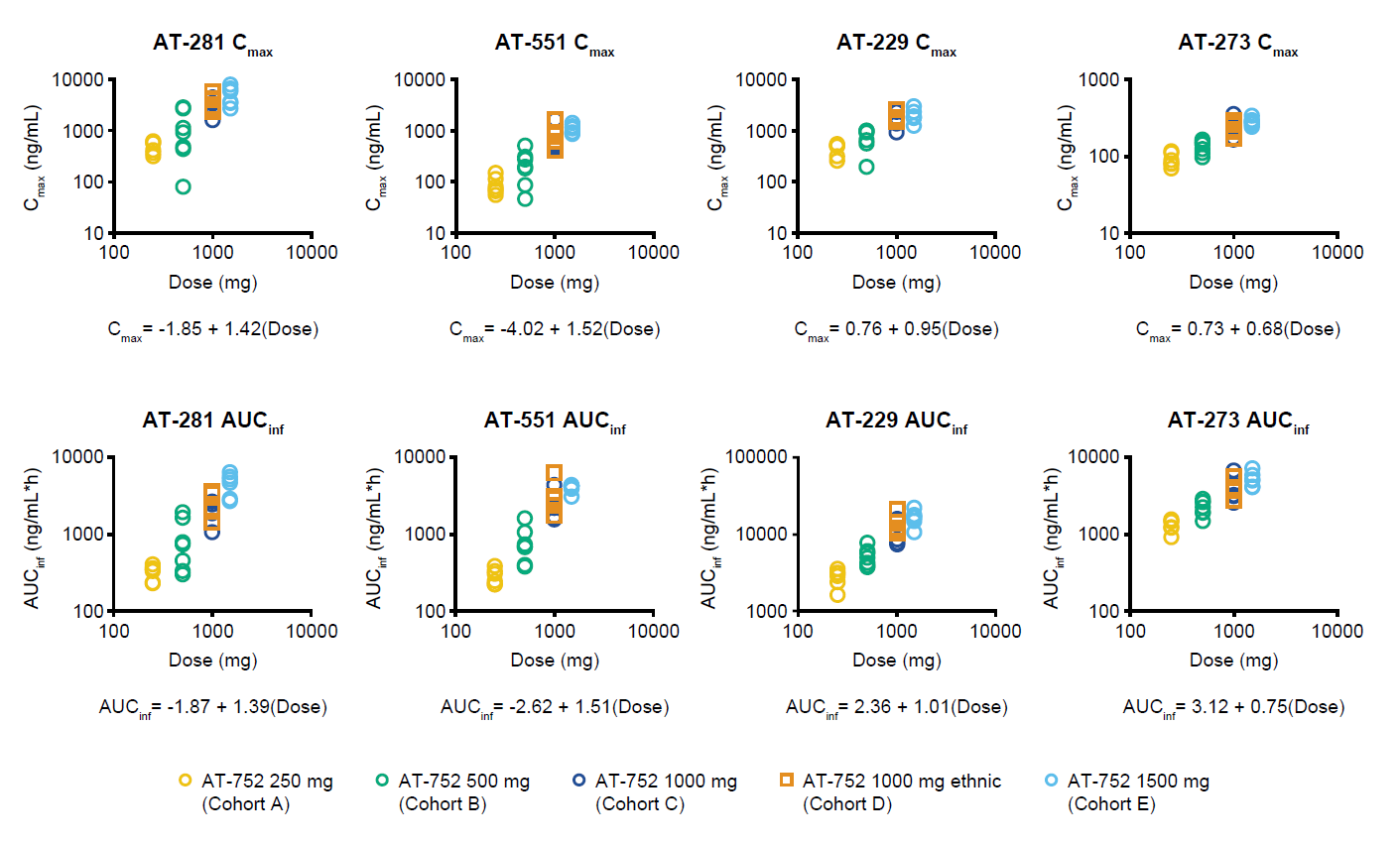
**Figure S1. Dose proportionality plots in SAD cohorts**

AUC_inf_, area under the curve extrapolated to infinity; C_max_, maximum plasma concentration; SAD, single ascending dose.
